# An implementation study of the service model for genetic risk-based stratified breast cancer screening – Estonian results of the BRIGHT project

**DOI:** 10.1101/2024.10.22.24315198

**Authors:** Madli Tamm, Peeter Padrik, Kristiina Ojamaa, Anette Paas, Anni Lepland, Krista Kruuv-Käo, Liis Leitsalu, Siim Sõber, Laura Roht, Sander Pajusalu, Tiina Kahre, Anna Padrik, Jagnar Pindmaa, Kadri Luga, Ly Rootslane, Anne Ilves, Sulev Ulp, Kersti Kallak, Ave-Triin Tihamäe, Neeme Tõnisson

## Abstract

Breast cancer (BC) remains the most common malignant tumor site and the leading cause of cancer-related deaths in women despite the wide availability of screening programs and personalized treatment options. The BRIGHT study tested a genetic risk-based personalized BC screening service model in women younger than 50 years, using telemedicine and home-based testing. Participants underwent polygenic risk score and monogenic pathogenic variant testing. This type of screening model demonstrated feasibility, clinical utility, and acceptability. It has the potential to enhance BC screening programs, particularly for younger women and those at higher genetic risk, while avoiding unnecessary interventions for low-risk individuals.

## Background

Breast cancer (BC) remains the most prevalent cancer among women and the leading cause of cancer deaths globally, representing a significant public health challenge. Every year adds 2.3 million new diagnoses and more than 660,000 deaths worldwide (1). Mammography screening has been shown to reduce BC mortality by 20–47% overall (2–5). The starting age for BC screening has been chosen to achieve an optimal balance between BC frequency, detection rates, clinical benefits, cost-effectiveness, and potential harms (false positives, overdiagnosis, and minimal radiation exposure) (6). In many guidelines, biennial screening for women aged 50–69 has been considered to be the most optimal (7,8). However, 29% of BC cases are diagnosed in women under the age of 50 (9), a demographic group that standard screening programs in Europe typically do not cover. Because it is impractical to screen all younger women, and biennial screening for high-risk women over 50 may miss interval cases, it is unreasonable to implement uniform screening for all age groups (10,11). These limitations underscore the need for a more nuanced approach to screening that can account for individual risk factors beyond age alone.

Stratified personalized screening and clinical recommendations to balance benefits, harms, and cost-effectiveness has long been envisioned as a more efficient model that aligns screening resources with those at elevated risk, thus potentially improving outcomes through earlier detection while simultaneously reducing unnecessary interventions for those at low risk, especially among younger women. Reasons why personalized screening has not been widely implemented include the need for a practical, applicable risk assessment model in routine practice, the absence of a cost-effective organizational model for risk-stratified screening, and unclear attitudes towards it from both women and medical professionals. (12–15)

In addition to gender and age, genetic predisposition is a crucial component of BC risk. Hereditary factors account for around one-third of the total BC risk (16) and form the basis of current models for stratified BC screening. Monogenic pathogenic variants (MPVs) in high- and moderate-risk cancer predisposition genes (e.g., *BRCA1*, *BRACA2*, *CHEK2*) have effects significant enough to warrant monogenic testing (17–19). However, only a small fraction (4–10%) of BC cases are caused by known MPVs (18,20), and MPV testing is typically conducted based on the fulfillment of high-risk criteria only (21). While MPV testing has been widely used in healthcare for some time, the polygenic risk score (PRS), another significant component of genetic predisposition, has predominantly been used only in research (22,23). The PRS is the combined effect of individual BC susceptibility single nucleotide polymorphisms (SNPs) identified by genome-wide association studies (24,25). Although individual SNPs may confer only modest disease risk, the combined impact of all BC-associated SNPs on risk can be substantial. A considerable portion (more than 30%) of BC risk variation is explained by BC-associated SNPs (11,26–28). Additionally, PRS can address a broader range of women regardless of their family cancer history (29).

The consideration of genetic risk must be included in national screening programs because clinical recommendations will not be accurate without them. Simulations suggest that risk-based preventive activities could provide cost savings and health benefits (30,31), and there is growing evidence base for the use of PRS in personalized BC screening (11). However, consensus clinical models for implementing PRS in routine, personalized BC screening have yet to be developed, and the optimal service model for personalized BC screening and its real applicability and feasibility as a systemic service remains unclear (13,23,32).

Our aim in the current “Be RIGHT with breast cancer risk management” (BRIGHT) study was to test a genetic risk-based personalized BC screening service model in real-world healthcare using different participant inclusion channels, PRS testing, and a family cancer history questionnaire to determine the need for MPV testing.

## Methods

### Recruitment and data/sample collection

The Estonian arm of the BRIGHT study was performed in 2022 and recruited 800 healthy women aged 35–49 who had not otherwise been invited into population BC screening. Participation was not dependent on valid health insurance. Women with already diagnosed malignancies or hereditary cancer syndromes, those already tested for MPVs and PRSs, or those with Ashkenazi Jewish ethnicity were excluded from this study. The general study scheme is shown in Figure 1.

**Figure 1.**
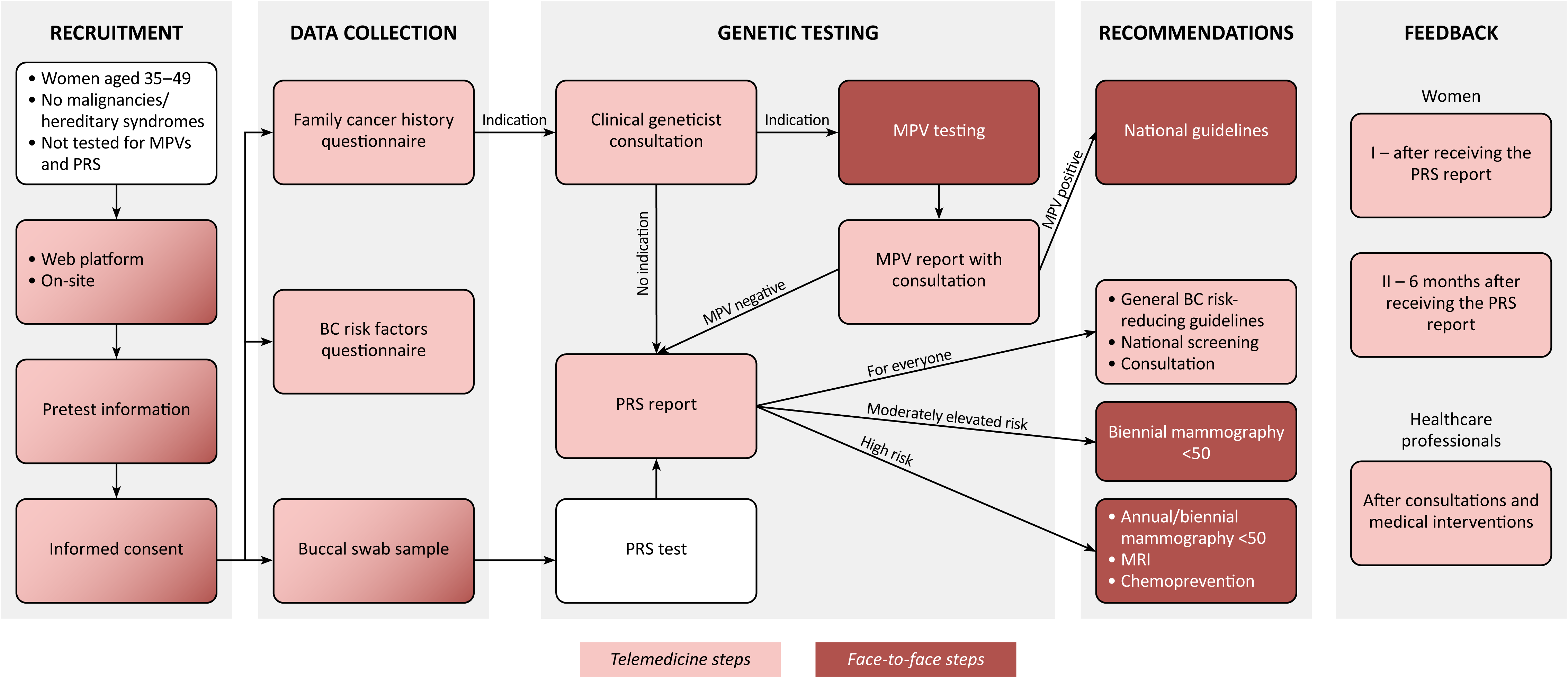
The BRIGHT study scheme for the Estonian arm. BC: breast cancer; PRS: polygenic risk score; MPV: monogenic pathogenic variant; MRI: magnetic resonance imaging.

Four different channels were used to recruit women for the study. There were opportunities to join the study by visiting a web portal, the Breast Clinic at Tartu University Hospital, participating pharmacies, or the primary healthcare center. Women got pretest information either digitally on the web portal or from a healthcare professional at the recruitment site. Informed consent forms for the study and PRS analysis were signed digitally in the web portal or on paper at the recruitment sites. DNA collection for the PRS testing was performed using a non-invasive buccal swab sample (AnteBC test, OÜ Antegenes, Estonia), either as a home test using parcel machines to receive and send the sample kit or at the participating sites. For data protection, buccal swab samples were pseudonymized by a research nurse.

Participants also filled out the questionnaires about family cancer history (modified from the hereditary breast and ovarian cancer guidelines of Tartu University Hospital) and BC clinical risk factors (see Additional file 1). If a participant used the web portal to enter the study, she filled out the questionnaires herself. If a participant enrolled at the hospital, pharmacy, or primary healthcare center, questionnaires were filled out with the help of investigators. Participant age distribution was assessed using chi-squared and binomial tests. All data collected from the participants is given in Additional file 2.

### PRS testing

PRS testing was performed for all women from buccal swab samples using the AnteBC test (OÜ Antegenes, Estonia). It is registered as an in vitro diagnostic device in the EUDAMED database (UDI-DI: 04745010362019), in the Estonian Medical Devices Database (EMDDB code: 14726), and in the UK MHRA Registry (GMDN code: 59918) for assessment of women’s polygenic risk of developing BC. The test combines information from 2803 BC-related genetic variants in an additive model described by Padrik et al. (33).

Based on the results of the AnteBC test, a report was generated with information about the PRS along with relative and absolute risk estimates, derived from individual and population average 10-year BC risks according to the iCARE model by Choudhury et al. (34). The PRS report was made available to participants and healthcare professionals in two telemedicine platforms - the Antegenes’ portal and the Estonian e-Health Record System. An example of the PRS report is shown in Additional file 3.

### Clinical interventions based on polygenic risk

All study participants received written PRS reports with post-test information and clinical recommendations along with general guidelines for reducing the risk of BC. All women were offered an opportunity to receive additional post-test consultations either online with a study nurse or face-to-face with an oncologist at Tartu University Hospital.

There are no international guidelines for BC screening recommendations based on PRS testing results. Therefore, recommendations from a previously published paper (33) were discussed with the Estonian expert advisory board of the BRIGHT study, and clinical recommendations for primary and secondary prevention of BC at different polygenic risk levels were established.

Participants were divided into four categories for recommendations based on their relative lifetime risks (RR) according to the PRS results: risk class 1 - up to average (RR < 1), risk class 2 - moderately elevated (1 ≤ RR < 2), risk class 3 - high (2 ≤ RR < 3), or risk class 4 - very high (RR ≥ 3) (Table 1). Recommendations within each risk class were further individualized according to a 10-year absolute risk estimate of 1.51%, corresponding to the average absolute BC risk in Estonia at the age of 50 (33). Women were recommended to begin biennial mammography screening at the age of reaching baseline risk, annual mammography screening at the age of reaching 2x baseline risk (3.02%), and magnetic resonance imaging screening at the age of reaching 3x baseline risk (4.53%). Participants received their PRS test results as PRS percentile, not risk group. If the participant’s age exceeded the recommended screening starting age, the patient was recommended to begin screening at their current age. To enter the mammography screening program, women received a digital referral note.

**Table 1.** Recommendations for personalized BC screening based on PRS results.

| **Risk class** | **Risk category** | **Description** | **Clinical recommendations** |
| --- | --- | --- | --- |
| 1 | Risk not elevated | Relative risk is up to the population's average | National standard screening recommendation. In Estonia, biennial mammography screening starting from the age of 50 |
| 2 | Moderately elevated risk | Relative risk increases up to 2-fold | Biennial mammography screening initiation at the age of attaining a 10-year risk equivalent to that of a genetically average 50-year-old* |
| 3 | High risk | Relative risk increases 2- to 3-fold | Biennial mammography screening initiation at the age of attaining a 10-year risk equivalent to that of a genetically average 50-year-old* |
|  |  |  | Annual mammography screening initiation starting from an age where 10-year risk attains 2-fold that of a genetically average 50-year-old* |
|  |  |  | Discussion about the usage of BC risk-reducing hormonal chemoprevention (tamoxifen, aromatase inhibitors) with a medical professional |
| 4 | Very high risk | Relative risk increases more than 3-fold | Biennial mammography screening initiation at the age of attaining a 10-year risk equivalent to that of a genetically average 50-year-old* |
|  |  |  | Annual mammography screening initiation starting from an age where 10-year risk attains 2-fold that of a genetically average 50-year-old* |
|  |  |  | Annual or biennial magnetic resonance imaging starting from an age where 10-year risk attains 3-fold that of a genetically average 50-year-old* |
|  |  |  | Discussion about the usage of BC risk-reducing hormonal chemoprevention (tamoxifen, aromatase inhibitors) with a medical professional |

\* If the recommended age was below the participant's current age, starting at the current age was recommended.

### MPV testing

Based on the family cancer history questionnaire, participants with suspected hereditary cancer cases among their first- and second-degree relatives received a digital referral note for counseling with a clinical geneticist. The specialist then decided whether MPV testing was indicated or not. MPV testing was done using Illumina’s TruSight Hereditary Cancer Panel (Illumina, Inc., San Diego, CA, USA), covering 113 genes related to various hereditary cancers, including all known genes related to hereditary BC. MPV testing was carried out in the Genetics and Personalized Medicine Clinic, Tartu University Hospital, and was initiated from the participants or the closest available cancer cases among their relatives. When an MPV was known in the family, only that specific exon was tested in participants. Cascade screening was initiated when an MPV was detected in a participant’s sample.

All participants who received MPV testing also underwent a second genetic counseling via telemedicine to discuss test results. The participant with an MPV was referred to further follow-up interventions according to relevant national guidelines. When giving clinical recommendations, both MPV and PRS results were considered.

### Feedback questionnaires

Acceptability and preferences regarding personalized prevention were investigated using questionnaires for study participants and medical personnel. Feedback was collected digitally from all participants 4–6 weeks after receiving the PRS report (I – short-term) and again 6–9 months after receiving the PRS report (II – long-term). Participants who had not responded to the first questionnaire could still respond to the second questionnaire. All healthcare professionals participating in this study were asked to complete the feedback questionnaire after all participants had received their results, consultations, and medical interventions. Feedback questionnaires were developed based on questionnaires and findings from previous studies (35–39) and are provided in Additional file 4. Questionnaire responses were anonymous, and results are reported as summary statistics.

Participant questionnaires mainly consisted of multiple-choice questions on a five-point Likert scale with an option to comment on some topics in their own words. The first participant feedback questionnaire included questions about the clarity of information provided during the study, difficulties with family cancer history collection, emotional responses to receiving the PRS report, and user experience with the digital portal. The second participant feedback questionnaire included questions about long-term psychological responses, decision regret, coping and perceived control, and with whom they shared the results.

The healthcare professionals’ feedback questionnaire was divided into five different topics: study preparation, recruitment, post-test consultation, study database, and BC PRS test. The questionnaire mainly consisted of multiple-choice answers with the option to comment on every topic in their own words and included questions about how they assessed the study activities and their attitudes toward personalized genetic risk assessment.

### Data management

All documents were stored securely in a study-specific database, accessible only to authorized study staff. Individual-level data protection complied with the General Data Protection Regulation (40) and relevant national legislative acts. The personal data of study participants was pseudonymized as soon as they entered the study database, and the pseudonymization key will be deleted at the end of the study. Data quality was monitored by the BRIGHT study team.

## Results

### Participant enrollment and characteristics

After the public announcement of the study, all four recruitment channels were popular and were filled up within a few days of opening the appointment times, with web portal and pharmacies being the first to be fully utilized. All channels recruited similar numbers of women with similar median age.

One woman was excluded from the study as she did not answer either the family cancer history or BC risk factors questionnaires. As a result, the study cohort comprised 799 Estonian women. The baseline characteristics of the 799 enrolled women are summarized in Table 2. The median age at the time of recruitment was 40 years (range 35–49). There were statistically more women (*P* = 2.567×10^−7^) in the younger age group and fewer women (*P* = 6.947×10^−12^) in the age group that would start their national BC screening program within the next five years. All study participants were of European descent. Based on the family cancer history questionnaire, 105 (13.1%) women had either breast or prostate cancer in a first- or second-degree relative.

**Table 2.** Baseline characteristics of the Estonian BRIGHT study cohort (*n* = 799).

| <u>Recruitment channels</u> | *n* (%) | Median age (range) |
| --- | --- | --- |
| Web portal | 192 (24.0) | 41 (35–49) |
| Pharmacy | 199 (24.9) | 41 (35–49) |
| Breast clinic | 207 (25.9) | 40 (35–49) |
| Family physician | 201 (25.2) | 39 (35–49) |

| <u>Distribution of the age groups</u> | *n* (%) | *P*-value |
| --- | --- | --- |
| 35–39 | 337 (42.2) | $2.567 \times 10^{-7}$ |
| 40–44 | 284 (35.5) | 0.2023 |
| 45–49 | 178 (22.3) | $6.947 \times 10^{-12}$ |

| <u>Breast cancer in relatives*</u> | *n* (%) |
| --- | --- |
| Under the age of 51 | 63 (7.9) |
| Breast or prostate cancer | 105 (13.1) |

**Non-genetic risk factors**
| <u>BMI**</u> | *n* (%) | Median BMI (range) |
| --- | --- | --- |
| <18.5: underweight | 28 (3.5) | 18.1 (17.3–18.4) |
| 18.5–24.9: normal weight | 448 (56.1) | 21.9 (18.5–24.9) |
| 25.0–29.9: overweight | 207 (25.9) | 26.9 (25.0–29.8) |
| ≥30.0: obesity | 116 (14.5) | 33.1 (30.0–46.8) |
| <u>Menopause</u> |  |  |
| Yes | 19 (2.4) |  |
| No | 775 (97.0) |  |
| Prefer not to say | 5 (0.6) |  |
| <u>Previous pregnancy</u> |  |  |
| Yes | 739 (92.5) |  |
| No | 58 (7.3) |  |
| Prefer not to say | 2 (0.3) |  |
| <u>Contraceptive pill</u> |  |  |
| Yes | 662 (82.9) |  |
| No | 137 (17.1) |  |
| <u>Alcohol consumption</u> |  |  |
| Yes | 632 (79.1) |  |
| No | 157 (19.6) |  |
| Prefer not to say | 10 (1.3) |  |

\* Reported by the participant in the questionnaire
\*\* Body mass index categories according to the World Health Organization

The study cohort was similar to the general population of Estonian women of the same age. Among the cohort, 56.1% had normal weight and 40.4% were overweight. A majority of participants reported current or previous use of alcohol (79.1%) or contraceptive pills (82.9%). Most of the women (92.5%) had been pregnant and 2.4% had gone through menopause.

### PRS testing

The distribution of participants into risk groups based on the PRS is shown in Table 3. Most women (58.7%) were in risk class 1, meaning they had no elevated polygenic risk of having BC. A total of 330 (41.3%) women had elevated BC risk, and three women had very high risk.

**Table 3.** Women’s distribution into risk groups based on the PRS results.

| **Risk class** | ***n* (%)** | **Median age (range)** | **Median percentile (range)** | **Median PRS (range)** | **Median RR (range)** | **Median AR (range)** |
| --- | --- | --- | --- | --- | --- | --- |
| 1 | 469<br>(58.7) | 40 (35–49) | 22 (1–50) | -0.79<br>(-3.36–0.00) | 0.68<br>(0.19–1.00) | 0.60<br>(0.15–1.45) |
| 2 | 282<br>(35.3) | 40 (35–49) | 70 (51–92) | 0.52<br>(0.00–1.39) | 1.29<br>(1.00–1.96) | 1.23<br>(0.55–2.73) |
| 3 | 45<br>(5.6) | 40 (35–49) | 97 (93–99) | 1.75<br>(1.44–2.24) | 2.30<br>(2.00–2.94) | 2.15<br>(1.18–4.49) |
| 4 | 3<br>(0.4) | Ages between the range of 35–49 | 100 (99–100) | 2.54<br>(2.31–3.01) | 3.33<br>(3.02–4.15) | 4.32<br>(3.17–6.55) |

*PRS: polygenic risk score expressed as standard deviation; RR: relative lifetime risk; AR: absolute 10-year risk expressed as a percentage*

During the year of participation in the study, 124 (15.5%) women with a median age 44 (range 35– 49) already had the same or higher 10-year absolute BC risk levels (range 1.51–6.55%, with a median of 1.90%) compared to the average absolute BC risk in Estonia at the age of 50 (1.51%).

### Clinical interventions based on polygenic risk

Women with no elevated BC risk (*n* = 469; 58.7%) received a recommendation to begin biennial mammography screening at the age of 50, which is the national standard in Estonia at the moment. Of the women with an elevated polygenic risk (*n* = 330), a total of 307 received recommendation to begin screening before the age of 50. Among them, 124 (40.4%) received recommendation and a referral note to begin mammography screening in the current year. For the remaining 23 women the 10-year absolute BC risk level was calculated to be ≥1.51% when they turned 50 years old; these women were thus advised to begin screening at age 50.

Of the 124 referred women, 93 (75.0%) had mammograms taken by the time the study database was closed. The clinical findings of these women are summarized in Table 4. Briefly, 77 (82.8%) had no findings, 14 (15.0%) had benign findings, one (1.1%) had a precancerous lesion, and one (1.1%) had carcinoma in situ. Women with a precancerous lesion and a stage 0 cancer diagnosis had no family cancer history. The woman with carcinoma in situ was also MPV negative based on the test ordered after the cancer diagnosis.

**Table 4.** Clinical findings for women who initiated screening based on their elevated PRS results.

| Clinical finding | *n* (%) | Median age (range) | Median RR (range) |
| --- | --- | --- | --- |
| Normal | 77 (82.8) | 44 (35–49) | 1.82 (1.11–4.15) |
| Benign finding | 14 (15.0) | 46 (38–49) | 1.72 (1.12–2.91) |
| Precancerous lesion | 1 (1.1) | Age between the range of 45–49 | 1.83 |
| Carcinoma in situ | 1 (1.1) | Age between the range of 45–49 | 1.60 |

*RR: relative lifetime risk*

### Post-test polygenic risk consultation

After receiving the PRS results, 90 (11.3%) out of the 799 women registered for the post-test appointment. The consultation was done via telemedicine with a study nurse (*n* = 54; 60.0%) or face-to-face with an oncologist (*n* = 36; 40.0%). Based on the enrollment method, the numbers of women who completed a post-test consultation were as follows: web portal 23 (25.6%), breast clinic 27 (30.0%), pharmacy 21 (23.3%), and primary healthcare center 19 (21.1%). All MPV-negative women got a post-test polygenic risk consultation during their appointment with a clinical geneticist. The majority of women who registered for post-test consultation had an elevated polygenic risk (*n* = 63, 70.0%). Of the 124 women who should have begun screening due to elevated polygenetic risk, 28 (22.6%) completed a post-test consultation. And of the 183 women who should begin screening before the age of 50, 32 (17.5%) completed a post-test consultation.

### MPV testing

Based on the self-administered family cancer history questionnaire, 152 (19.0%) women were referred to a clinical geneticist and 99 (65.1%) attended the appointment. Of these, 92 (92.9%) women or their family members were referred to MPV testing and 90 (97.8%) completed the testing. Two (2.2%) women did not wish to proceed with the MPV testing. Based on the clinical geneticist consultation, 7 (7.1%) women were not tested for MPVs because the family history was insufficient.

Four (4.4%) of the women tested had MPV-positive test results. A detailed overview of the MPV findings and the clinical recommendations given are shown in Table 5. Three women had lower polygenic BC risk compared to the Estonian population average and, therefore, began screening based on the MPV finding. One woman also had elevated polygenic risk (PRS RR = 2.19), and she was advised to begin biennial BC screening within three years.

**Table 5.** Details about the MPV findings and recommendations given to the women.

| Partici-<br>pants'<br>age* | MPV finding | Pathogenicity** | Fami-<br>lial | Clinical recommendation<br>based on the MPV | PRS<br>RR |
| --- | --- | --- | --- | --- | --- |
| 40–44 | NM_000057.4(BLM):<br>c.1642C>T p.(Gln548*) | Pathogenic | N/A | Annual mammography | 0.79 |
| 35–39 | NM_001128425.1<br>(MUTYH):c.536A>G<br>p.(Tyr179Cys) | Pathogenic | Yes | Body awareness, regular<br>gynecological<br>examinations, and national<br>BC screening | 2.19 |
| 45–49 | NM_007194.4(CHEK2):<br>c.319+2T>A p.? | Pathogenic /<br>Likely pathogenic | Yes | Annual mammography,<br>consider colonoscopy from<br>the age of 50 at 7-year<br>intervals, or yearly fecal<br>occult blood testing | 0.71 |
| 40–44 | NM_000535.7(PMS2):<br>c.1588C>T, p.(Gln530*) | Likely pathogenic | N/A | Annual mammography,<br>colonoscopy at 1–3-year<br>intervals, consider<br>prophylactic acetylsalicylic<br>acid treatment and<br>gastroscopy every 2 to 4<br>years. Annual<br>gynecological examination<br>for endometrial cancer<br>prevention. | 1.00 |

\*Participants' age is between the given age range
\*\*Pathogenicity based on ClinVar (41)
MPV: monogenic pathogenic variant; PRS: polygenic risk score; RR: relative lifetime risk

### Participants’ feedback

Feedback questionnaires were sent to all 799 participants; 240 (30.0%) responded to the first questionnaire and 255 (31.9%) to the second. Of the 255 respondents to the second questionnaire, 235 (92.2%) reported that they had responded to both questionnaires. Women reported receiving information about the study mainly from a friend/family member, social media, or a healthcare professional. 95% of the participants were satisfied with the telemedicine solution. The most problematic for them was the report comprehensibility (*n* = 26; 10.8%).

Feedback about participants’ emotional responses to receiving the PRS report, how they rated the information received about genetic risk, and their coping and perceived control are shown in Figure 2. Most women reported positive sentiments, found the information useful, and that they were able to cope with the knowledge about their polygenetic risk.

**Figure 2.**
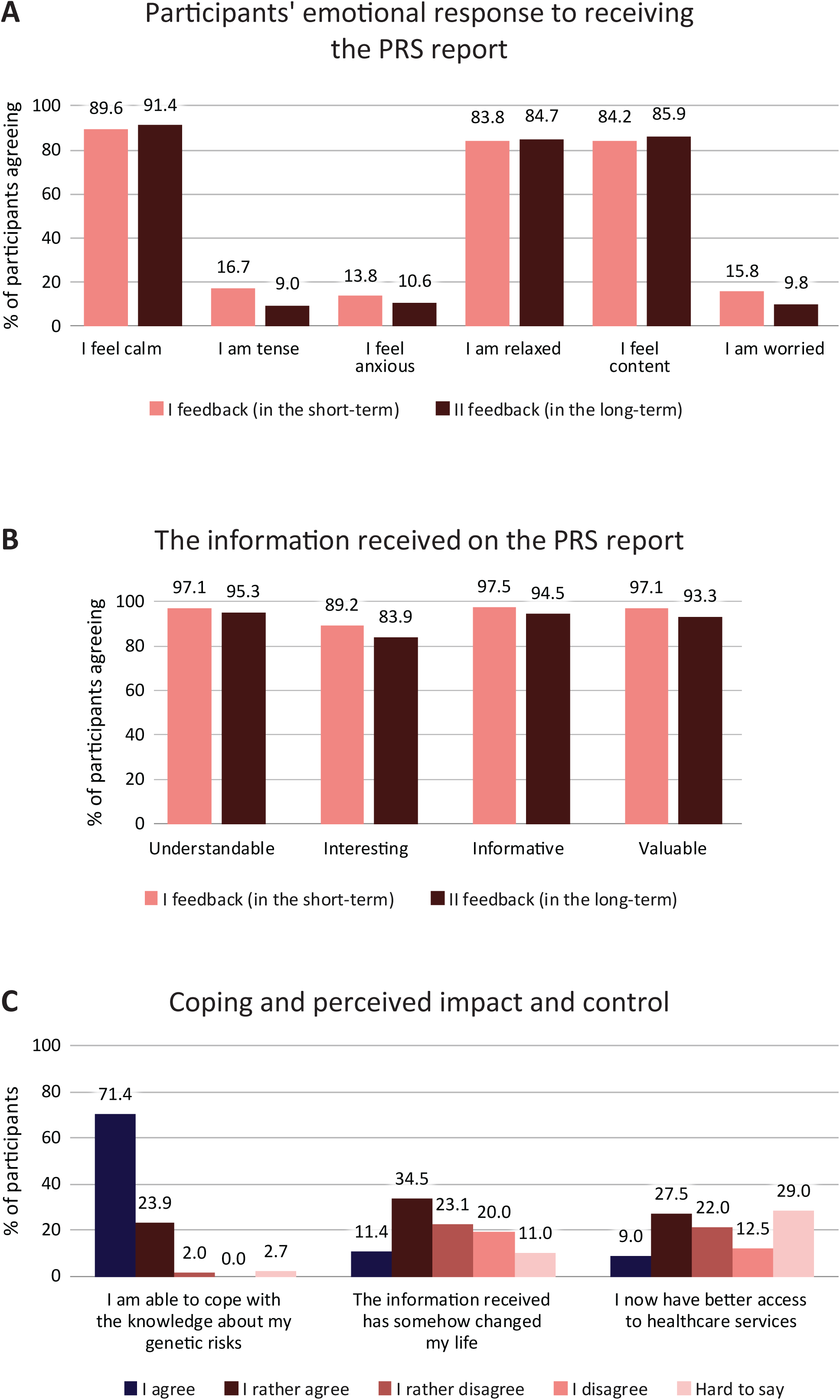
Feedback about participants’ (a) short- and long-term emotional responses to receiving the PRS report, (b) thoughts about the information received, and (c) level of coping and perceived impact and control.

The information about the study was generally considered sufficient, clear, and understandable. A large majority of participants (83.8%) reported that they had no difficulties giving answers about family cancer history. Women reported feeling comfortable with resources for further medical action (61.7%) and that they intended to follow the recommendations provided to them (94.1%). Some women (33.8%) reported needing further consultation about their polygenic risk results. Most of the women who received the post-test consultation rated it useful (92.5%).

A great majority of women were glad they had the opportunity to participate in the BRIGHT study (99.6%). They considered it a right and wise decision and would make the same choice again. Most women shared their experiences as part of the BRIGHT study with a family member (79.6%) or a friend (62.0%). Half of the women (53.3%) reported not having looked for additional information and the other half looked for information from the internet (33.7%) or a healthcare professional (25.9%).

Eight (3.3%) women in the first feedback and 13 (5.1%) in the second feedback asked for additional information. Their questions were mainly about how to notice changes in their breasts and how to get a referral for mammography in the future. There were also several comments about the lack of appointment times with the clinical geneticist and the need for a more simplified risk report. We gave written responses using their provided contact information. Two women did not provide any contact information and could not be responded to.

### Healthcare professionals’ feedback

Feedback questionnaires for healthcare professionals were sent to 32 individuals, and 18 (56.3%) responded. Many (38.9%) of them had not previously heard of PRS testing. Still, they considered the information provided in the pre-study training sufficient to carry out the BRIGHT study, and they rated nothing to be difficult to explain to the participants during enrollment. Four (22.2%) had already used PRS information in their clinical practice. Of the 13 healthcare professionals who used the doctors’ portal during the study, 11 (84.6%) were very satisfied with it.

Seven healthcare professionals who answered the feedback questionnaire were involved in the recruitment of participants. Most healthcare professionals (85.7%) agreed or rather agreed that the time scheduled for the enrollment was adequate. Feedback about the categories of questions asked by participants to healthcare professionals and aspects considered difficult by healthcare professionals during enrollment are shown in Figure 3. The main questions asked by participants were about the time taken to obtain results (85.7%), the definition of cancer sites (71.4%), and prohibited activities immediately prior to the buccal swab test (71.4%). Healthcare professionals considered it difficult to take family cancer history (57.1%) and explain risk results (42.9%) during enrollment. There was one healthcare professional who rated nothing to be difficult during enrollment.

**Figure 3.**
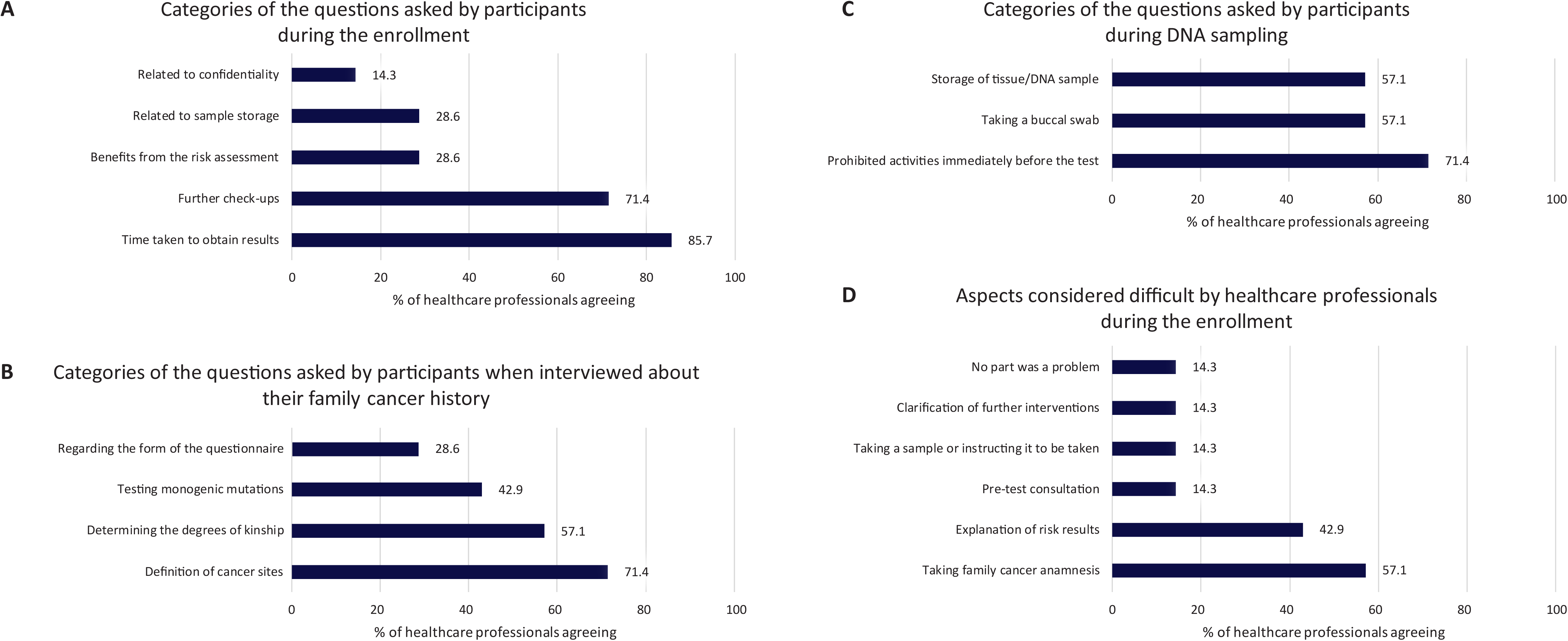
Summary of healthcare professionals’ feedback about questions asked by participants (a) during enrollment, (b) when interviewed about their family cancer history, and (c) during DNA sampling. Also assessed were (d) aspects considered difficult by healthcare professionals during enrollment.

All healthcare professionals who gave post-test consultations (*n* = 9) rated the PRS report as manageable, with the most challenging parts being the explanation of absolute vs. relative risk (55.6%) and monogenic vs. polygenic risk (55.6%). About half of the respondents (55.5%) would appreciate additional information to carry out the post-test consultation. The average time used for the consultation was 15–30 minutes. Healthcare professionals reported most participants feeling calm and relaxed (77.8%), with initially worried or anxious participants being more relieved by the end of the consultation (88.9%). During the post-test consultation, based on healthcare professionals’ feedback, participants asked mainly where to turn for further follow-up check-ups in the following years (77.8%) and how they could get a referral (77.8%).

Most respondents (88.9%) considered genetic risk assessment important in BC prevention, and most (83.3%) would consider using BC PRS testing in their clinical practice. Nurses reported that they do not have enough time to integrate new services in the near future; on the other hand, pharmacists were eager to be involved in the screening service. Some healthcare professionals recommended giving a more simplified PRS report to the women, and some were concerned that PRS results may provide false security or, on the contrary, raise anxiety. They also mentioned that they would not use PRS testing in their clinical practice until international guidelines were in place.

## Discussion

Measures to prevent premature deaths from BC should encompass prevention, early detection, and improved treatment. Whereas treatments are already somewhat personalized, prevention should also consider a woman’s personal risk of developing BC (42). The current BC screening programs are suboptimal because these consider only age as a risk factor, thereby excluding younger women with elevated risk and not providing optimal screening intervals for high-risk older women (43). Combined risk assessment models such as CanRisk (44–46) provide more accurate BC risk assessments, but their complexity and cost make them impractical for routine use in healthcare systems (47). Therefore, more simplified and practical genetic risk-based approaches, such as those using PRS, offer a pragmatic and effective alternative for improving BC screening programs.

The BRIGHT study evaluated the implementation of a risk-tailored BC screening service model in younger women currently excluded from the standard national screening program. This model calculated genetic predisposition via PRS testing for all women and recommended MPV testing based on a family cancer history questionnaire. The study also aimed to enhance BC prevention by raising awareness of BC risk factors and early symptoms. To our knowledge, the BRIGHT project is the first to demonstrate a fully digital personalized BC screening service model with home-based PRS testing.

To assess feasibility, our study used various channels (telemedicine platform, breast clinic, general practitioner practice, and pharmacies) to recruit participants. The effective recruitment through these diverse channels suggests that integrating multiple and accessible options can enhance participation rates in national screening programs, particularly among younger women who may not be reached by traditional enrollment methods.

The current standard approach for assessing genetic risks includes pre- and post-test genetic counseling (48). However, this approach is not feasible for population-based screening due to insufficient resources within healthcare systems and among healthcare professionals (49,50). Therefore, we replaced traditional counseling with written information provided before and after PRS testing while offering the option of post-test verbal consultation upon request. All women who underwent MPV testing received pre- and post-test counseling by a clinical geneticist. However, for MPV-negative women verbal counseling is not necessarily required, as supported by existing literature (51). The present results suggest that traditional face-to-face counseling may be unnecessary for PRS testing, as most women did not opt for post-test consultation while still reporting high satisfaction rates. Specifically, only about a tenth of women used the verbal post-test consultation option, indicating that written and digital communications can effectively support genetic risk assessment, aligning with findings from other studies (52,53). By leveraging written information and digital tools, our model addresses the limitations of resource-intensive traditional counseling. This approach can enhance the feasibility of implementing genetic risk-based screening in broader populations, ensuring that high-risk individuals receive appropriate preventive measures while maintaining efficiency and cost-effectiveness.

BC PRS testing is currently implemented in Estonian healthcare outside of the national BC screening program. For PRS to be effectively applied in clinical practice, it must be complemented by clinical recommendations for preventive activities tailored to different levels of risk (13,54,55). To this end, we have developed personalized recommendations based on relative risks compared to individuals of the same nationality, age, and sex, as well as estimated absolute risks.

Genetic risk information provides a more precise screening approach, and PRS is a strong independent risk component (11,29,56). A genetic risk-based approach to BC screening is feasible for several reasons. Genetic predispositions do not fluctuate over time, making this a stable and reliable risk assessment method. Additionally, PRS testing can be conducted at home, making it convenient and accessible for a wide population. This approach also allows for the stratification of women into different risk categories, enabling more personalized screening schedules and interventions. This stratification is particularly important as it can help avoid unnecessary interventions for low-risk individuals while ensuring that high-risk individuals receive the appropriate level of screening and preventive measures. In our study, two out of five women (41.3%) had elevated BC risk and were recommended to a personalized screening program. By identifying individuals at higher genetic risk earlier, it is possible to begin screening before the age of 50 and align more closely with their individual risk profiles. It is important to note that the women with a precancerous lesion and a stage 0 cancer diagnosis did not have any family cancer indication for earlier screening and presumably would have been diagnosed with later-stage cancer after symptoms had appeared. Based on the participants’ feedback, the given personal clinical recommendations were understandable and women intended to follow them.

The presence of an MPV is a significant risk factor for BC, necessitating personalized prevention. The currently accepted standard is to perform MPV testing on predominantly healthy women based on family cancer history. However, this approach is not perfect and leaves a significant portion of MPV carriers undetected (37,57). The high cost of testing currently prevents the implementation of MPV testing for all women. Systematic testing, where the target group is determined by a questionnaire, has not been implemented in national BC screening programs. In the BRIGHT study, we tested the implementation of a questionnaire-based decision-making process. Based on the small proportion of “do not know” answers and women’s feedback about the knowledge of their family cancer history, we can hypothesize that decisions made based on the questionnaire were accurate. We had only 4.4% MPV-positive women out of all the participants tested, and none of them had the usual BC high-risk variants. We assume this is probably because not all women (34.9%) with familiar BC indications attended the clinical geneticist appointment, thus we had a relatively small MPV cohort (*n* = 90), and the overall prevalence of the pathogenic *BRCA* variants in Estonia is very low (<1%) (37). Nevertheless, we do recommend implementing questionnaire-based MPV testing in the decision-making process for national screening programs.

This study had several limitations. First, we had enrollment bias. We did not control the age distribution during enrollment, and during analysis, we noticed that younger women were more eager to learn about their BC risk. Also, we can assume that enrolled women were more health conscious. Second, the study was not designed to assess randomized long-term effects, which could have enabled analysis of real changes in the incidence and mortality of BC in the study cohort. The provided personalized medicine interventions were preventive, and the pilot project lasted less than 2.5 years. Thus, the effects on mortality could not be assessed. Third, because the feedback questionnaire was voluntary, it had a relatively low response rate. Fourth, MPV testing was done only for a subset of participants and the testing was voluntary. Finally, this study did not consider breast density, gynecological differences, previous treatment history, or lifestyle factors, and did not combine PRS and MPVs to determine a woman’s overall risk. Women received their polygenic and monogenic BC risk reports separately, and the genetic risk factors were not combined with the non-genetic ones. We chose not to use combined risk models due to their limited use in healthcare, the complexity of data collection, and the difficulty of integrating them into our pipeline. Instead, we focused on PRS testing and familial cancer history for MPV testing, which proved to be a more feasible and practical approach.

After participating in the BRIGHT study, women were more aware of genetic and non-genetic BC risk factors, knew their personal polygenic BC risk, and were informed about how to better prevent or detect BC. Younger women with elevated risk were able to participate in an intensified BC screening program, presumably increasing their survival rate when affected by the disease. At the same time, unnecessary screening of lower-risk women was avoided. The study did not involve de-escalated screening recommendations that deviated from the current standard of care according to national guidelines. All participants with lower-than-average estimated risk were recommended to begin standard BC screening at age 50.

The BRIGHT study resulted in an easy-to-perform personalized BC screening service model that can be applied in other countries worldwide. Our study assessed that this type of personalized BC screening service is acceptable, feasible, has clinical utility, and healthcare systems are ready to implement it. The large interest in joining the study, successful implementation, and positive feedback suggest that most BC screening services could be provided digitally. Such BC screening services will save time and medical personnel resources. We envisage that in the near future, all BC screening participants could give a DNA sample during a regular check-up with a family doctor, the usual trip to the pharmacy, or even comfortably at home. In addition to personalized results, this new type of screening service model could include more women than the current national programs.

### Conclusions

The BRIGHT study demonstrated that a personalized BC screening service model based on genetic predisposition is feasible within the Estonian healthcare system. This model enables the inclusion of women with elevated genetic risk who have not yet reached the standard screening age. It is feasible to implement a service model that combines PRS assessments with the current standard practice of testing for MPVs based on family cancer history. To achieve broad inclusion of women in the personalized service model various engagement channels could be used. The study also indicated that in many cases, pre- and post-test counseling is not necessary when implementing PRS genetic testing; instead, comprehensive written information about the test’s nature and results, along with written clinical recommendations, is often sufficient. Telemedicine service models and genetic home testing can reduce the burden on the healthcare system. This enhanced BC screening service model could ultimately improve BC prevention and early detection by making it more accessible and precise.

In conclusion, the genetic risk-based personalized BC screening service model in real-world healthcare is feasible and acceptable for participating women as well as for medical professionals.

## Abbreviations

BC: breast cancer
BRIGHT: Be RIGHT with breast cancer risk management
MPV: monogenic pathogenic variant
PRS: polygenic risk score
RR: relative lifetime risk
SNP: single nucleotide polymorphism

## Declarations

### Ethics approval and consent to participate

The investigators ensured that this study was conducted according to the principles of the Declaration of Helsinki, with relevant regulations, and Good Clinical Practice. This study was approved by the Estonian Committee on Bioethics and Human Research (permission number 1.1-12/1930), the parallel study arms by the Portuguese Ethics Committee of the Lisbon Academic Medical Center (permission number 177/22), and the Swedish Ethical Review Authority (permission number 2022-03074-01). All study participants signed informed consent to participate in the study, and their samples were de-identified. The BRIGHT study is registered in the ISRCTN registry under ID ISRCTN29884654.

### Consent for publication

Not applicable

### Trial registration

The BRIGHT study is registered in the ISRCTN registry under ID ISRCTN29884654.

### Availability of data and materials

All materials used for this study are included in the main or supplementary information files. The participants’ datasets generated and analyzed during the current study are not publicly available due to sensitivity reasons but are available from the corresponding author upon reasonable request. Data are located in controlled access data storage at OÜ Antegenes.

### Competing interests

PP has ownership in OÜ Antegenes. AP, KKK, SS, AP, JP, and NT are receiving salaries from OÜ Antegenes. PP, KO, AL, LL, LR (Laura Roht), SP, TK, AI, SU, KK, ATT, and NT are receiving salaries from the Tartu University Hospital.

## Funding

This project was supported via the EIT Health BRIGHT innovation activity (project #230121). EIT Health is supported by the European Institute of Innovation and Technology (EIT), a body of the European Union. Views and opinions expressed are, however, those of the authors only and do not necessarily reflect those of the European Union or the EIT. Neither the European Union nor the granting authority can be held responsible for them.

SP received support from the Estonian Research Council grant PSG774.

## Authors’ contributions

PP and NT had the initial idea, designed and supervised the study. KKK, KO, AP (Anna Padrik), LR (Ly Rootslane), KL, SP, and SU participated in the study design.

AP (Anette Paas), KL, and AI enrolled and consulted participants and acquired the health data. SU acquired the mammography data. LR (Laura Roht) and NT conducted MPV counseling with participants. TK coordinated the MPV testing. KK and ATT participated in the post-test consultations and interpreted the participants’ clinical analysis results. JP and AP (Anna Padrik) managed the collected data. SS interpreted the collected data.

LL participated in designing the feedback questionnaires and substantively revised the feedback paragraphs of the manuscript. AL participated in designing the feedback questionnaire for healthcare professionals, acquired the feedback data, and proofread the manuscript.

MT contributed to the study design, performed data analysis and interpretation, and wrote the manuscript. PP helped to draft the manuscript. AP (Anette Paas), KKK, SS, and NT substantively revised the manuscript.

All authors read and approved the final manuscript.

## Data Availability

All materials used for this study are included in the main or supplementary information files. The participants datasets generated and analyzed during the current study are not publicly available due to reasons of sensitivity but are available from the corresponding author upon reasonable request. Data are located in controlled access data storage at OU Antegenes.

## Acknowledgments

The authors acknowledge all clinical and research colleagues who contributed to the sample collection and DNA analysis. Especially Mari Nelis and the Core Facility of Genomics, University of Tartu, and the Department of Laboratory Genetics, Genetics and Personalized Medicine Clinic, Tartu University Hospital. We want to thank all the participating women for donating their buccal swab samples and relevant clinical data. We thank Kätlin Eiche for coordinating and managing the BRIGHT project. Write Science Right provided professional proofreading.

## Additional Files

**Additional File 1** - Comma-separated values file “Questionnaires”, questions asked of participants about family cancer history and BC clinical risk factors

**Additional File 2** - Comma-separated values file “Data collected”, data collected about participants

**Additional File 3** – Portable document format file “PRS Report”, an example of the PRS report given to the participants

**Additional File 4** - Comma-separated values file “Feedback”, feedback questionnaires sent to the participants and healthcare professionals

## Notes

### Competing Interest Statement

PP has ownership in OU Antegenes. AP, KKK, SS, AP, JP, and NT are receiving salaries from OU Antegenes. PP, KO, AL, LL, LR (Laura Roht), SP, TK, AI, SU, KK, ATT, and NT are receiving salaries from the Tartu University Hospital.

### Clinical Trial

ISRCTN29884654

### Author Declarations

Ethics committee of the Estonian Committee on Bioethics and Human Research gave ethical approval for this work (permission number 1.1-12/1930). Ethics committee of the Portuguese Ethics Committee of the Lisbon Academic Medical Center gave ethical approval for this work (permission number 177/22). Ethics committee of the Swedish Ethical Review Authority gave ethical approval for this work (permission number 2022-03074-01).

## References

1. Bray F, Laversanne M, Sung H, Ferlay J, Siegel RL, Soerjomataram I, et al. Global cancer statistics 2022: GLOBOCAN estimates of incidence and mortality worldwide for 36 cancers in 185 countries. CA: A Cancer Journal for Clinicians. 2024;74(3):229–63.

2. Myers ER, Moorman P, Gierisch JM, Havrilesky LJ, Grimm LJ, Ghate S, et al. Benefits and Harms of Breast Cancer Screening: A Systematic Review. JAMA. 2015 Oct 20;314(15):1615–34.

3. Duffy SW, Vulkan D, Cuckle H, Parmar D, Sheikh S, Smith RA, et al. Effect of mammographic screening from age 40 years on breast cancer mortality (UK Age trial): final results of a randomised, controlled trial. Lancet Oncol. 2020 Sep;21(9):1165–72.

4. Duffy SW, Tabár L, Yen AMF, Dean PB, Smith RA, Jonsson H, et al. Mammography screening reduces rates of advanced and fatal breast cancers: Results in 549,091 women. Cancer. 2020 Jul 1;126(13):2971–9.

5. Tabár L, Dean PB, Chen TH, Yen AM, Chen SL, Fann JC, et al. The incidence of fatal breast cancer measures the increased effectiveness of therapy in women participating in mammography screening. Cancer. 2019 Feb 15;125(4):515–23.

6. World Health Organization. WHO Position Paper on Mammography Screening. Geneva: World Health Organization; 2014.

7. Ren W, Chen M, Qiao Y, Zhao F. Global guidelines for breast cancer screening: A systematic review. The Breast. 2022 Aug 1;64:85–99.

8. Katsika L, Boureka E, Kalogiannidis I, Tsakiridis I, Tirodimos I, Lallas K, et al. Screening for Breast Cancer: A Comparative Review of Guidelines. Life (Basel). 2024 Jun 19;14(6):777.

9. Global Cancer Observatory. Cancer (IARC) TIA for R on [Internet]. [cited 2024 Jun 27]. Available from: https://gco.iarc.fr/

10. Nelson HD, Pappas M, Cantor A, Griffin J, Daeges M, Humphrey L. Harms of Breast Cancer Screening: Systematic Review to Update the 2009 U.S. Preventive Services Task Force Recommendation. Ann Intern Med. 2016 Feb 16;164(4):256–67.

11. Mars N, Kerminen S, Tamlander M, Pirinen M, Jakkula E, Aaltonen K, et al. Comprehensive Inherited Risk Estimation for Risk-Based Breast Cancer Screening in Women. JCO. 2024 May;42(13):1477–87.

12. Shieh Y, Eklund M, Sawaya GF, Black WC, Kramer BS, Esserman LJ. Population-based screening for cancer: hope and hype. Nat Rev Clin Oncol. 2016 Sep;13(9):550–65.

13. Pashayan N, Antoniou AC, Ivanus U, Esserman LJ, Easton DF, French D, et al. Personalized early detection and prevention of breast cancer: ENVISION consensus statement. Nat Rev Clin Oncol. 2020 Nov;17(11):687–705.

14. Schmutzler RK, Schmitz-Luhn B, Borisch B, Devilee P, Eccles D, Hall P, et al. Risk-Adjusted Cancer Screening and Prevention (RiskAP): Complementing Screening for Early Disease Detection by a Learning Screening Based on Risk Factors. Breast Care. 2021 Aug 12;17(2):208–23.

15. Laza C, Niño de Guzmán E, Gea M, Plazas M, Posso M, Rué M, et al. “For and against” factors influencing participation in personalized breast cancer screening programs: a qualitative systematic review until March 2022. Archives of Public Health. 2024 Feb 22;82(1):23.

16. Mucci LA, Hjelmborg JB, Harris JR, Czene K, Havelick DJ, Scheike T, et al. Familial Risk and Heritability of Cancer Among Twins in Nordic Countries. JAMA. 2016 Jan 5;315(1):68–76.

17. Breast Cancer Association Consortium, Dorling L, Carvalho S, Allen J, González-Neira A, Luccarini C, et al. Breast Cancer Risk Genes - Association Analysis in More than 113,000 Women. N Engl J Med. 2021 Feb 4;384(5):428–39.

18. Slavin TP, Maxwell KN, Lilyquist J, Vijai J, Neuhausen SL, Hart SN, et al. The contribution of pathogenic variants in breast cancer susceptibility genes to familial breast cancer risk. NPJ Breast Cancer. 2017 Jun 9;3:22.

19. Lee K, Seifert BA, Shimelis H, Ghosh R, Crowley SB, Carter NJ, et al. Clinical validity assessment of genes frequently tested on hereditary breast and ovarian cancer susceptibility sequencing panels. Genetics in Medicine. 2019 Jul 1;21(7):1497–506.

20. Tung N, Battelli C, Allen B, Kaldate R, Bhatnagar S, Bowles K, et al. Frequency of mutations in individuals with breast cancer referred for BRCA1 and BRCA2 testing using next-generation sequencing with a 25-gene panel. Cancer. 2015 Jan 1;121(1):25–33.

21. Sessa C, Balmaña J, Bober SL, Cardoso MJ, Colombo N, Curigliano G, et al. Risk reduction and screening of cancer in hereditary breast-ovarian cancer syndromes: ESMO Clinical Practice Guideline. Annals of Oncology. 2023 Jan 1;34(1):33–47.

22. Khera AV, Chaffin M, Aragam KG, Haas ME, Roselli C, Choi SH, et al. Genome-wide polygenic scores for common diseases identify individuals with risk equivalent to monogenic mutations. Nat Genet. 2018 Sep;50(9):1219–24.

23. Lewis CM, Vassos E. Polygenic risk scores: from research tools to clinical instruments. Genome Medicine. 2020 May 18;12(1):44.

24. Yang X, Kar S, Antoniou AC, Pharoah PDP. Polygenic scores in cancer. Nat Rev Cancer. 2023 Sep;23(9):619–30.

25. Choi SW, Mak TSH, O’Reilly PF. Tutorial: a guide to performing polygenic risk score analyses. Nat Protoc. 2020 Sep;15(9):2759–72.

26. Mavaddat N, Michailidou K, Dennis J, Lush M, Fachal L, Lee A, et al. Polygenic Risk Scores for Prediction of Breast Cancer and Breast Cancer Subtypes. The American Journal of Human Genetics. 2019 Jan 3;104(1):21–34.

27. Jia G, Lu Y, Wen W, Long J, Liu Y, Tao R, et al. Evaluating the Utility of Polygenic Risk Scores in Identifying High-Risk Individuals for Eight Common Cancers. JNCI Cancer Spectrum. 2020 Jun 1;4(3):pkaa021.

28. Mavaddat N, Pharoah PDP, Michailidou K, Tyrer J, Brook MN, Bolla MK, et al. Prediction of breast cancer risk based on profiling with common genetic variants. J Natl Cancer Inst. 2015 May;107(5):djv036.

29. Wolfson M, Gribble S, Pashayan N, Easton DF, Antoniou AC, Lee A, et al. Potential of polygenic risk scores for improving population estimates of women’s breast cancer genetic risks. Genet Med. 2021 Nov;23(11):2114–21.

30. Pashayan N, Morris S, Gilbert FJ, Pharoah PDP. Cost-effectiveness and Benefit-to-Harm Ratio of Risk-Stratified Screening for Breast Cancer: A Life-Table Model. JAMA Oncology. 2018 Nov 1;4(11):1504–10.

31. Maas P, Barrdahl M, Joshi AD, Auer PL, Gaudet MM, Milne RL, et al. Breast Cancer Risk From Modifiable and Nonmodifiable Risk Factors Among White Women in the United States. JAMA Oncology. 2016 Oct 1;2(10):1295–302.

32. Kumuthini J, Zick B, Balasopoulou A, Chalikiopoulou C, Dandara C, El-Kamah G, et al. The clinical utility of polygenic risk scores in genomic medicine practices: a systematic review. Hum Genet. 2022;141(11):1697–704.

33. Padrik P, Puustusmaa M, Tõnisson N, Kolk B, Saar R, Padrik A, et al. Implementation of Risk-Stratified Breast Cancer Prevention With a Polygenic Risk Score Test in Clinical Practice. Breast Cancer(Auckl). 2023 Jan 1;17:11782234231205700.

34. Choudhury PP, Maas P, Wilcox A, Wheeler W, Brook M, Check D, et al. iCARE: An R package to build, validate and apply absolute risk models. PLOS ONE. 2020 veebr;15(2):e0228198.

35. Marteau TM, Bekker H. The development of a six-item short-form of the state scale of the Spielberger State—Trait Anxiety Inventory (STAI). British Journal of Clinical Psychology. 1992;31(3):301–6.

36. Brehaut JC, O’Connor AM, Wood TJ, Hack TF, Siminoff L, Gordon E, et al. Validation of a Decision Regret Scale. Med Decis Making. 2003 Jul 1;23(4):281–92.

37. Leitsalu L, Palover M, Sikka TT, Reigo A, Kals M, Pärn K, et al. Genotype-first approach to the detection of hereditary breast and ovarian cancer risk, and effects of risk disclosure to biobank participants. Eur J Hum Genet. 2021 Mar;29(3):471–81.

38. Jürgens H, Roht L, Leitsalu L, Nõukas M, Palover M, Nikopensius T, et al. Precise, Genotype-First Breast Cancer Prevention: Experience With Transferring Monogenic Findings From a Population Biobank to the Clinical Setting. Front Genet [Internet]. 2022 Jul 22 [cited 2024 Sep 23];13. Available from: https://www.frontiersin.org/journals/genetics/articles/10.3389/fgene.2022.881100/full

39. Leitsalu L, Reigo A, Palover M, Nikopensius T, Läll K, Krebs K, et al. Lessons learned during the process of reporting individual genomic results to participants of a population-based biobank. Eur J Hum Genet. 2023 Sep;31(9):1048–56.

40. Union PO of the E. Publications Office of the EU. Publications Office of the European Union; 2016 [cited 2024 Sep 21]. Regulation (EU) 2016/679 of the European Parliament and of the Council of 27 April 2016 on the protection of natural persons with regard to the processing of personal data and on the free movement of such data, and repealing Directive 95/46/EC (General Data Protection Regulation) (Text with EEA relevance). Available from: https://op.europa.eu/en/publication-detail/-/publication/3e485e15-11bd-11e6-ba9a-01aa75ed71a1/language-en

41. Landrum MJ, Lee JM, Riley GR, Jang W, Rubinstein WS, Church DM, et al. ClinVar: public archive of relationships among sequence variation and human phenotype. Nucleic Acids Res. 2014 Jan;42(Database issue):D980–985.

42. Roberts E, Howell S, Evans DG. Polygenic risk scores and breast cancer risk prediction. The Breast. 2023 Feb 1;67:71–7.

43. Mars N, Koskela JT, Ripatti P, Kiiskinen TTJ, Havulinna AS, Lindbohm JV, et al. Polygenic and clinical risk scores and their impact on age at onset and prediction of cardiometabolic diseases and common cancers. Nat Med. 2020 Apr;26(4):549–57.

44. Lee A, Mavaddat N, Wilcox AN, Cunningham AP, Carver T, Hartley S, et al. BOADICEA: a comprehensive breast cancer risk prediction model incorporating genetic and nongenetic risk factors. Genetics in Medicine. 2019 Aug 1;21(8):1708–18.

45. Carver T, Hartley S, Lee A, Cunningham AP, Archer S, Babb de Villiers C, et al. CanRisk Tool-A Web Interface for the Prediction of Breast and Ovarian Cancer Risk and the Likelihood of Carrying Genetic Pathogenic Variants. Cancer Epidemiol Biomarkers Prev. 2021 Mar;30(3):469–73.

46. Archer S, Villiers CB de, Scheibl F, Carver T, Hartley S, Lee A, et al. Evaluating clinician acceptability of the prototype CanRisk tool for predicting risk of breast and ovarian cancer: A multi-methods study. PLOS ONE. 2020 märts;15(3):e0229999.

47. Tsoulaki O, Tischkowitz M, Antoniou AC, Musgrave H, Rea G, Gandhi A, et al. Joint ABS-UKCGG-CanGene-CanVar consensus regarding the use of CanRisk in clinical practice. Br J Cancer. 2024 Jun;130(12):2027–36.

48. Berliner JL, Cummings SA, Boldt Burnett B, Ricker CN. Risk assessment and genetic counseling for hereditary breast and ovarian cancer syndromes-Practice resource of the National Society of Genetic Counselors. J Genet Couns. 2021 Apr;30(2):342–60.

49. K S, S K, Sa C. The past, present and future of service delivery in genetic counseling: Keeping up in the era of precision medicine. American journal of medical genetics Part C, Seminars in medical genetics [Internet]. 2018 Mar [cited 2024 Oct 10];178(1). Available from: https://pubmed.ncbi.nlm.nih.gov/29512888/

50. Bamshad MJ, Magoulas PL, Dent KM. Genetic counselors on the frontline of precision health. American Journal of Medical Genetics Part C: Seminars in Medical Genetics. 2018;178(1):5–9.

51. Swisher EM, Rayes N, Bowen D, Peterson CB, Norquist BM, Coffin T, et al. Remotely Delivered Cancer Genetic Testing in the Making Genetic Testing Accessible (MAGENTA) Trial: A Randomized Clinical Trial. JAMA Oncology. 2023 Nov 1;9(11):1547–55.

52. Marjonen H, Marttila M, Paajanen T, Vornanen M, Brunfeldt M, Joensuu A, et al. A Web Portal for Communicating Polygenic Risk Score Results for Health Care Use—The P5 Study. Front Genet. 2021 Oct 29;12:763159.

53. Biesecker BB, Lewis KL, Umstead KL, Johnston JJ, Turbitt E, Fishler KP, et al. Web Platform vs In-Person Genetic Counselor for Return of Carrier Results From Exome Sequencing: A Randomized Clinical Trial. JAMA Internal Medicine. 2018 Mar 1;178(3):338–46.

54. Shah PD. Polygenic Risk Scores for Breast Cancer-Can They Deliver on the Promise of Precision Medicine? JAMA Netw Open. 2021 Aug 2;4(8):e2119333.

55. Yanes T, Young MA, Meiser B, James PA. Clinical applications of polygenic breast cancer risk: a critical review and perspectives of an emerging field. Breast Cancer Research. 2020 Feb 17;22(1):21.

56. Zhang YD, Hurson AN, Zhang H, Choudhury PP, Easton DF, Milne RL, et al. Assessment of polygenic architecture and risk prediction based on common variants across fourteen cancers. Nat Commun. 2020 Jul 3;11:3353.

57. Grindedal EM, Heramb C, Karsrud I, Ariansen SL, Mæhle L, Undlien DE, et al. Current guidelines for BRCA testing of breast cancer patients are insufficient to detect all mutation carriers. BMC Cancer. 2017 Jun 21;17(1):438.

