## Additional file 3 for "An implementation study of the service model for genetic risk-based stratified breast cancer screening – Estonian results of the BRIGHT project"

### AnteBC

#### A polygenic risk score test for breast cancer

| | |
|---|---|
| Last name: | Kask |
| First name: | Mari |
| National ID: | 1234567890 |
| Age: | 33 |
| Nationality: | Estonian |
| Sample material: | buccal swab |
| Genotyping array: | Illumina Global Screening Array-24 Kit |
| Analysis and interpretation: | OÜ Antegenes (Licence L05386) |

#### Summary

The patient's polygenic risk score of breast cancer is 2.15 standard deviation (SD) units.

The result shows that the breast cancer polygenic risk score is 2.15 standard deviation units higher than the population average placing patient's risk score among 33-year-old women in the 99th percentile. **Meaning that more than 98% of women have lower and more than 1% of women have higher polygenic risk score.**

The patient's risk of developing breast cancer in the next 10 years is 1.29%. The 10-year average risk of developing breast cancer among 33-year-old women in Estonia is 0.44%. **In terms of relative comparison, the current result implies that the risk of developing breast cancer in the next 10 years is 2.96 times higher than the 10-year genetic risk among 33-year-old women on average.**

**Based on the breast cancer polygenic risk score test results, we recommend:**

- Mammography screening every two years starting at the age of 35.
- Mammography screening every year starting at the age of 42.
- Discuss the use of breast cancer risk decreasing hormonal chemoprevention (tamoxifen, aromatase inhibitors) with your doctor.
- Follow general guidelines to reduce the risk of breast cancer (see our recommendations).

The individual cancer risk can be high, whether or not there is a family history of cancer. Knowing the personal breast cancer risk is important because further clinical analysis can now be implemented according to the patient's risk. This way we can prevent the disease or detect it as early as possible.

### AnteBC test results and explanatory information

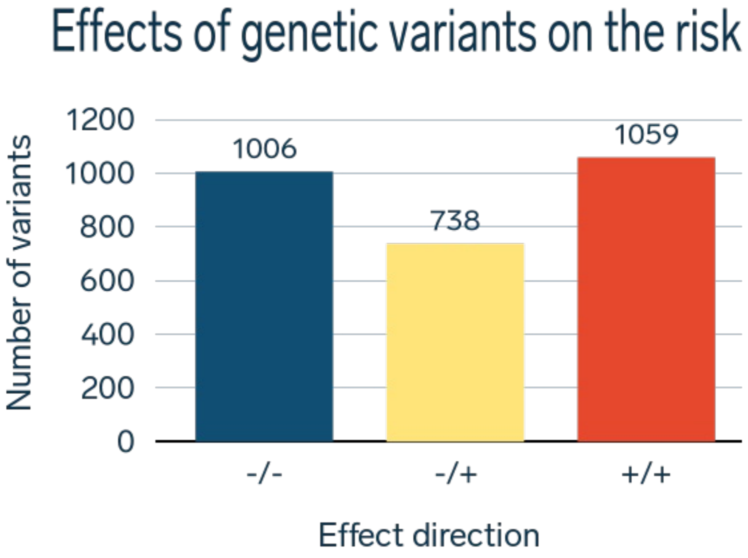


A typical human genome contains 23 pairs of chromosomes, for a total of 46 varying widely in size. In total, the genome contains around 6.4 billion different positions. The human genome has two genetic variants in each position (except variants on male sex chromosomes): one from the mother and the other from the father.

The AnteBC test includes a total of 2803 positions. Out of these, the patient has 1006 (-/-) positions in her genome where both variants reduce the risk. However, there are 738 (-/+) positions where one genetic variant increases, and the other decreases the risk, and 1059 (+/+) positions where both variants increase risk.

By analyzing all risk positions in the patient’s genome, we estimated that the patient’s risk score for developing breast cancer is 2.15 SD units. The risk score is higher than in 98% and lower than in 1% of 33-year-old women. In other words, the patient’s breast cancer risk score is placed in the 99th percentile of 33-year-old women.

### Patient and the general population

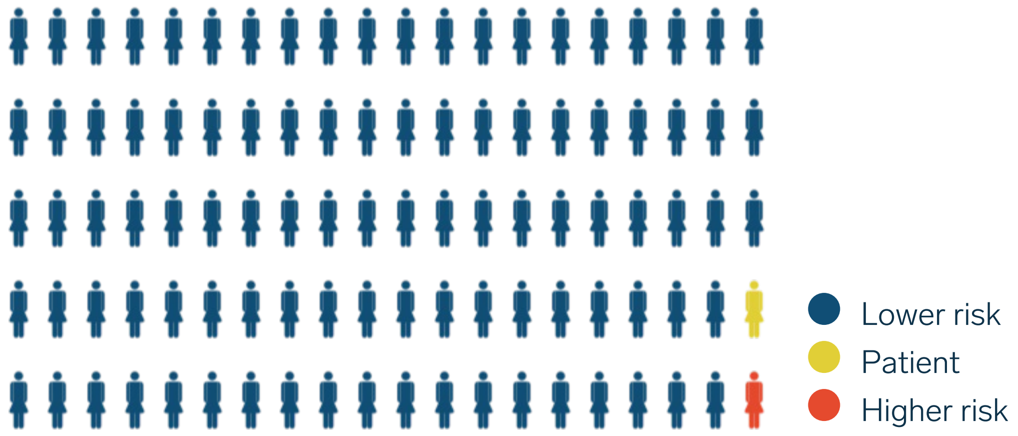


AnteBC test considers the patient's nationality, gender, age, and the demographic background of breast cancer. The patient's risk of developing breast cancer within the next 10 years is 1.29% (1.51–1.09%). About 129 women out of 10,000 will develop the disease. At the same time, the risk of breast cancer among 33-year-old women in Estonia is 0.44% (0.41–0.46%) meaning that the expected rate of developing the disease is 44 women out of 10 000.

#### 10-year risk of developing the disease

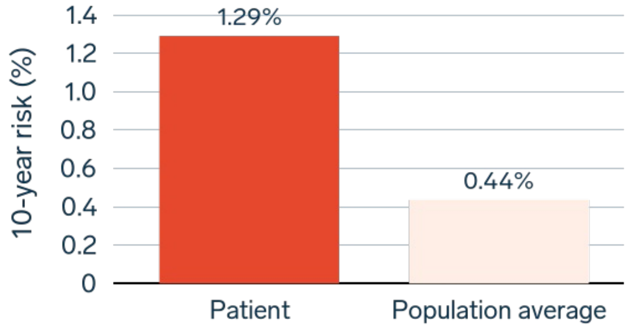


#### Population risk levels

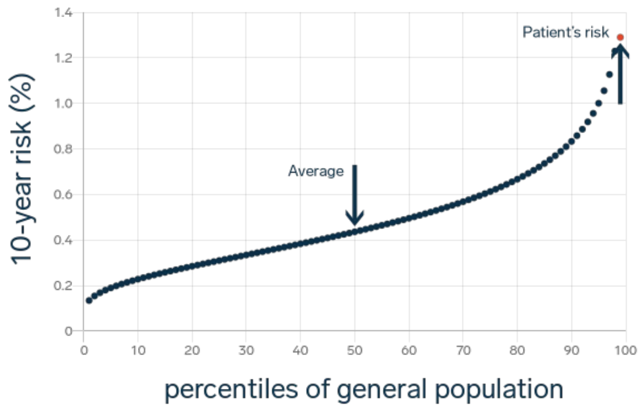

### AnteBC test clinical recommendations

The World Health Organization (WHO) recommends mammography screening for women between the ages of 50 and 69.

This guideline is also adopted in Estonia where women between the ages of 50 and 69 get screened every two years. Independent research has shown that high-quality mammography screening reduces the risk of breast cancer mortality by an average of 20% in this age group.

**Based on the AnteBC test, the patient's risk of developing breast cancer compared to genetically average 33-year-old women is 2.96 times higher.**

Based on the breast cancer polygenic risk score test results, we recommend:

- Mammography screening every two years starting at the age of 35.
- Mammography screening every year starting at the age of 42.
- Discuss the use of breast cancer risk decreasing hormonal chemoprevention (tamoxifen, aromatase inhibitors) with your doctor.
- Follow general guidelines to reduce the risk of breast cancer (see our recommendations).

In addition to the polygenic component used by the AnteBC test, there are also other breast cancer risk factors to be considered. Risk assessment could be affected by any previous breast diagnostic tests, occurrences of cancer in close (biological) relatives, or individual health behaviors.

#### Health behavior

- A body mass index greater than 30 increases the risk of breast cancer by a factor of 1.5 to 2. Thus, it is recommended for adults to maintain a body mass index between 18.5 and 24.9 or at least below 30.
- Physical activity is risk-reducing. At least 30 minutes of moderate-to-vigorous intensity physical activity is recommended on most days, a total of 1.5 to 4 hours per week.
- Consuming just one alcoholic drink a day increases breast cancer risk by 5%. Regular alcohol consumption should be avoided to reduce the risk of breast cancer.
- Using hormone replacement therapy (HRT) during menopause increases the risk of breast cancer. The risk of breast cancer increases with the use of estrogen and progestogen combination and estrogen alone. Therefore, the risk-benefit ratio of these drugs should be discussed with your doctor.
- Women who have never given birth and women who give birth to their first child at the age of 35 or older have a slightly higher risk of developing breast cancer.
- Smoking increases the risk of breast cancer.

### Body awareness

We recommend you to be aware of your body, including the condition and possible changes in your breasts. If you notice any of the symptoms listed below, we recommend that you seek medical attention. These may indicate the development of breast cancer:

- Abnormal changes in breast shape, size or color;
- New lump or mass in breast tissue;
- Pain or discomfort in one breast;
- Changes in the surface of the breast skin (looking like an orange peel), skin retraction, "wrinkling" or ulcer;
- Change of shape or position of a nipple or retraction;
- Bleeding or flushing around the nipple, discharge from the nipple;
- Enlargement of the axillary lymph nodes.

### AnteBC test general information

AnteBC is a genetic test that estimates a patient's risk of developing breast cancer. AnteBC test is based on the methodology of polygenic risk scores, which enables early detection and prevention of breast cancer.

In addition to the patient’s genetics, age, gender and ancestry, risk calculations also take into account the Estonia population average morbidity and mortality rates. As the risk of cancer increases with age, each patient is compared with people of the same age when evaluating the test results.

Genetic variants used in the AnteBC test are distributed throughout the genome. The AnteBC test includes a total of 2803 genetic variants that can increase or decrease the risk of breast cancer.

The result of the AnteBC is given as units of standard deviation (SD) that characterizes the patient's genetic risk compared to the population average taking into account patient’s ancestry (European, African, East Asian, South Asian or Mixed ancestry). For example, an outcome that exceeds 2.326 SD units corresponds to the highest level of risk in the 99th percentile. A result lower than -2.326 SD units corresponds to the lowest level of risk in the 1st percentile.

In case the patient's age exceeds the actual recommended starting age for screening or any other procedures, the report will state the patient's age for the start time.

### AnteBC test limitations

- AnteBC cannot be used to diagnose breast cancer.
- The risks identified by the AnteBC test take into account the polygenic risk, but do not consider other risk factors (see section Health behavior).
- An elevated risk estimated by the AnteBC test does not mean that the patient will develop breast cancer during their lifetime. Also, a moderate or low-risk score does not mean that the patient will not develop breast cancer.
- AnteBC test is patient-specific, it does not give any information about the risk of developing a disease in the patient’s family or close relatives, i.e. polygenic risk score-based disease risks may not be transmitted directly from parents to children.
- AnteBC test does not analyze rare risk increasing mutations in single genes, e.g., *BRCA1*, *BRCA2*, *CHEK2*, *PALB2*, *ATM*, *TP53*, *CDH1*, *STK11*, etc. Therefore, we recommend testing of rare risk increasing mutations in single genes if the following criteria are met:
    1. The patient has a history of breast, ovarian, fallopian tube or peritoneal cancer.
    2. A biological relative has a mutation in a single gene predisposed to breast cancer (*BRCA1*, *BRCA2*, etc.).
    3. A first- or second-degree biological relative has been diagnosed with breast

cancer (under the age of 45 years), pancreatic cancer, ovarian cancer, metastatic prostate cancer; two or more cases of breast cancer in one person or a first/second-degree male biological relative has a history of breast cancer.

4. Biological relatives have had three or more tumors associated with hereditary cancer syndromes.
5. Patient is of Ashkenazi Jewish origin.

- The AnteBC test is based on up-to-date scientific data. However, the field of genetics is constantly evolving which may lead to changes in the risk assessments in the future as additional information becomes available. Therefore, also the clinical recommendations based on the test results may change.
- Different polygenic risk score models of the same trait may give different estimates to the individual's risks due to differences in the genetic variants included in the model and their weights.
- The result of this test should be applied in context with other relevant clinical data. In addition to the possible genetic predisposition, other risk factors also affect the risk of developing breast cancer.

### AnteBC test genetic consultation

To better understand the results of the AnteBC test **an individual post-test consultation is recommended**. Consultation is conducted by healthcare professionals who ensure that the results are understandable and useful.

### Contact

OÜ Antegenes (Licence L05386)
Registry code: 14489312

www.antegenes.com
